# A rapid and sensitive Loop-Mediated Isothermal Amplification (LAMP) assay for detection of *mecA* antibiotic resistance gene in clinical and food bacterial isolates

**DOI:** 10.64898/2026.11.13.26367169

**Authors:** Léo Baldenweck, Mila Djisalov, Debora Pinamonti, Marisa Manzano, Ivana Gadjanski, Jasmina Vidic

## Abstract

An advanced LAMP assay targeting the *mecA* gene in methicillin-resistant *Staphylococcus aureus* (MRSA) was developed and validated using clinical and foodborne *S. aureus* isolates. Reference strains, including both methicillin-sensitive and methicillin-resistant *S. aureus*, were used to evaluate and optimize assay performance. With the newly designed ID16 LAMP primer set, the detection limit for *mecA* was 15 fg/µL of total genomic DNA, equivalent to 5 copies/µL, and the assay turnaround time was only 25 min. All *mecA*-positive isolates identified by conventional qPCR were successfully detected by the LAMP assay. Overall, the developed LAMP method provides a rapid, highly specific, and sensitive alternative for *mecA* detection.

## 1. Introduction

Antimicrobial resistance (AMR) represents a critical global health challenge, contributing to millions of deaths annually and imposing a substantial economic burden on healthcare systems and societies worldwide [1, 2]. Resistant bacteria are primarily transmitted through three major reservoirs: community-associated pathways (person-to-person spread), healthcare-associated environments (usually via contaminated surfaces and medical equipment), and livestock-associated routes (predominantly through food products of animal origin). In those reservoirs, AMR genes can be spread from resistant strains to sensitive ones through the horizontal gene transfer mechanisms such as conjugation, transduction, and transformation, often facilitated by mobile genetic elements like integrons and transposons [3]. Moreover, an extensive use of antibiotics will favour the emergence of resistance by creating a selective pressure environment[4].

*Staphylococcus aureus* is a Gram-positive bacterium, usually commensal, as part of the normal microbiota of the body and can be found in nares, skin, and the gastrointestinal tract [5]. However, *S. aureus* can be opportunistic pathogen leading to pneumonia, endocarditis, and sepsis, among others [6]. Furthermore, group of strains of *S. aureus* that are resistant to methicillin and other β-lactam antibiotics *i*.*e*., MRSA is one of the leading causes of healthcare-associated and community-acquired infections, according to WHO [7, 8]. The ability of MRSA to resist methicillin can be due to the *mecA* gene, present in a mobile genetic element, SCCmec [8, 9], which encodes for a modified methicillin-target, PBP2a, with reduced affinity for β-lactam antibiotics [10]. A high genetic flexibility of *S. aureus*, enables easy development of novel defence mechanisms, yielding to new multi-resistant strains.

To manage the spread of AMR, the WHO identifies four pillars: prevention of infections, better access to health services, appropriate treatment, and accurate and fast diagnosis [11]. Traditional antibiotic resistance diagnostics mainly involve the bacterial isolation and the performing of an Antibiotic Susceptibility Test. This test is an official culture-based method where sensitivity to a given antibiotic is evaluated by an inhibition zone on agar plate. However, this method is time-consuming, often taking three or more days needed for bacterial isolation and culture, and may fail, if the patient is already under an antibiotic treatment [12], or in the case that bacteria enter in viable but not cultivable (VBNC) state [13]. Consequently, traditional antibiotic tests delay targeted treatment and often necessitate initial use of broad-spectrum empirical therapies.

To limit the spread of MRSA and enable early administration of targeted antibiotics, the development of novel strategies for rapid and sensitive detection of resistant strains is urgently required. Genotypic testing for *mecA* is widely used to rapidly optimize antimicrobial therapy in *S. aureus* infections. Molecular-based techniques, such as Polymerase Chain Reaction (PCR), permit a faster detection, with response in less than a day. However, PCR technique has high costs, requires trained staff, specific machines, and remains constrained to a lab-setting [14].

To address these drawbacks of PCR, isothermal amplification methods have been developed to accurately detect nucleic acids through faster amplification and easier adaptation to point-of-care format [15], since there is no need for temperature variations and specific hardware components such as a thermal cycler. Loop-mediated isothermal amplification (LAMP) is one of the most promising isothermal techniques as it uses a set of 4 to 6 primers specific to the target gene and delivers results within less than 1 hour [16]. When coupled with a colorimetric kit incorporating the pH-sensitive dye (such as Phenol Red), LAMP allows a naked-eye detection of amplification results, with a pink solution for a negative result and yellow for a positive result.

Here we present a LAMP assay for the sensitive detection of *mecA* within 25 min. We designed and validated *in silico* novel LAMP primers targeting *mecA*. After experimental validation using well-characterized *S. aureus* strains with or without the target gene, we applied the LAMP assays to screen clinical and foodborne *S. aureus* strains (Fig. S1). The LAMP assay results were confirmed by the qPCR detection for the same gene, which corroborates efficiency and accuracy of the LAMP assay.

## 2. Experimental

### 2.1. Bacterial strain and culturing

Five well-characterized *S. aureus* strains were used for primer validation: USA300-JE2[17], LAC[18] and COL[19], which hold *mecA* gene and are methicillin-resistant (referred to as MRSA), and RN-R[20], and Newman[21] which are methicillin-susceptible (referred to as MSSA).

The isolation and identification of eight foodborne *S. aureus* strains from raw cow milk and pork meat, were previously described in detail [22]. Briefly, starting from 150 mL of milk and 100 g of meat samples homogenized 1:2 in physiological solution, 10 mL were serially diluted and plated on Baird-Parker Agar supplemented with Egg Yolk Tellurire Emulsion (Oxoid, Milan, Italy). After incubation at 37°C for 48 h, shiny, black colonies, with double halo were selected and streaked on Plate Count Agar (Oxoid, Milan, Italy) for purification. *S. aureus* identification was performed by PCR targeting the species-specific thermonuclease *nuc* gene [23].

Twenty-two clinical *S. aureus* strains were isolated from the blood from anonymized patient collected during sepsis diagnostic procedures and were kindly provided by Jean-Louis Herrmann (Hôpital Raymond Poincaré, Garches, France).

All strains were cultivated in Brain Heart Infusion (BHI) broth at 37°C.

### 2.2. DNA extraction

An overnight culture was centrifuged at 8000 g, 4°C, for 10 min, and washed with TES buffer (10 mM Tris-HCl, 100 mM NaCl, 1.0 mM EDTA, pH of 7.8). The bacterial pellet was lysed using FastPrep-24 (MP Biomedicals, France) with the following settings: 3 cycles of 20 sec at speed 4 m/sec, with 5 min rest between each cycle. The genomic DNA (gDNA) was purified with the DNeasy blood and tissue kit (Qiagen, France). Quantification of the gDNA was performed by NanoDrop2000 (ThermoScientific). From these concentrations, the number of copies of the gene was calculated using the website NEBioCalculator from Biolabs with the following formula:

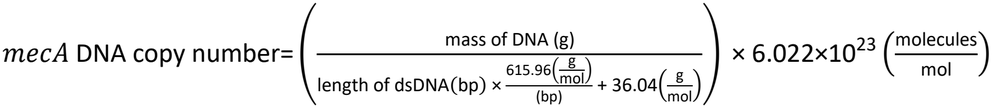

As *mecA* is a single-copy gene, we can approximate that one *mecA* DNA copy corresponds to one colony-forming unit (CFU).

### 2.4. LAMP primers design and in silico validation

The *mecA* gene sequence used for the LAMP primer design was extracted from the complete genome of *S. aureus* strain USA300JE2 (GenBank accession number NZ_CP020619), referred to as JE2. Prior to primer design, *mecA* sequence was analysed using the Nucleotide Basic Local Alignment Search Tool (BLAST) (https://blast.ncbi.nlm.nih.gov/Blast.cgi) to assess the level of conservation of the target regions across different bacterial strains. LAMP primer sets were designed using the PrimerExplorer V5 program (https://primerexplorer.eiken.co.jp/lampv5e/index.html), targeting different sequences within the specific regions of the *mecA* gene. To minimize the risk of undesired secondary structures, such as hairpins, homodimers, and heterodimers, the primer sequences were analysed using the OligoAnalyzer software provided by Integrated DNA Technologies (IDT), available at https://eu.idtdna.com/pages/tools/oligoanalyzer. Designed LAMP sequences and locations on the *mecA* sequence are showed in Supplementary data (Table S1 and Fig. S2).

Thirty-eight sequences of *Staphylococcus spp. mecA* were extract from NCBI (https://www.ncbi.nlm.nih.gov/) with the following search: PBP2a family beta-lactam-resistant peptidoglycan transpeptidase MecA. The sequences were aligned to highlight mismatch using Benchling (https://benchling.com). The accession number of the sequences are provided in Table S2.

### 2.4. PCR primer design and assays

The qPCR primers were designed using Primer3 from Benchling. The specificity to the target was verified by BLAST, and the absence of secondary structure was confirmed using ViennaRNA from Benchling. For the amplification of *mecA*, the following primers were used: mecA3: 5’ CCAATTCCACATTGTTTCGGTCTAA 3’[24] and qmecAR: 5’ CCAGGAATGCAGAAAGACCAAAGCA 3’. Primers were first validated by PCR using FIREPol DNA Polymerase Kit (Solisbiodyne, France). The reaction mix was composed of the FIREPol buffer BD X1, 2.5 mM MgCl_2_, 0.2 mM dNTP, 0.3 µM of each primer, 0.05 U/µL of FIREPol, for a total volume of 20 µL. The thermocycler Nexus GX2 (Eppendorf, France) was programmed for a first denaturation step of 95°C for 4 min, then 35 cycles of denaturation at 95°C for 17 sec, annealing at 60°C for 35 sec and extension at 72°C for 1 min. Then a final extension was performed at 72°C for 10 min.

qPCR was performed using the Luna Universal qPCR master mix from New England Biolabs (France). The reaction mix was composed of qPCR buffer X1, 0.3 µM of each primer for a total volume of 20 µL. The reaction was performed using qPCR StepOne Plus (Applied Biosystem, France) which was programmed for a first denaturation step of 95°C for 20 sec, then 40 cycles of denaturation at 95°C for 3 sec, annealing and extension at 60°C for 30 sec. A final step includes a denaturation of 15 sec and an annealing/extension of 1 min. A melting curve was then performed to check the specificity by increasing 0.5°C every 15 sec until reaching 95°C.

### 2.5. LAMP reaction

The LAMP reactions were performed with WarmStart LAMP Kit (DNA & RNA) for the Fluorimetric LAMP reaction, and with WarmStart Colorimetric LAMP 1X Master Mix (DNA & RNA) for colorimetric LAMP reaction (NewEngland Biolabs, France). Each reaction mixture contained 20 μL of reagents, including WarmStart LAMP 1x Master Mix, 1x LAMP primer mix (16 μM FIP and BIP, 2 μM F3 and B3, and 4 μM of the LF and LB), fluorescent dye 50X for fluorometric assays, 2 μL of DNA sample and nuclease-free water. Reactions were performed using the StepOne Plus system. All detection assays were performed in duplicate (technical replicates), and ultrapure water and gDNA of methicillin-sensitive strains were used as templates in negative controls.

### 2.6. Agarose gel electrophoresis

Products of the amplification were validated by loading in 2% agarose gel with ethidium bromide solution (Invitrogen, France) and running in electrophoresis unit Mupid One (Eurogentec) for 30 min at 135 V. The gel is then visualized with E-Box Gel documentation (Vilber, France).

### Detection of mecA in biological matrices

Adult bovine serum (CliniSense, France) was selected as model biological matrix and was used without further processing. *S. aureus* JE2 and RN-R cells from overnight BHI cultures were harvested, washed three times with TES buffer, and used to spike the two matrices. Bacterial cells were incubated in these media under gentle agitation at room temperature for 1 h. Cells were then lysed using FastPrep. The LAMP reaction was performed using 2 µL of clarify lysates.

### 2.8. Methicillin susceptibility tests

Agar spot diffusion method was performed to test susceptibility of *S. aureus* isolates to methicillin. *S. aureus* cells from an overnight BHI liquid culture were spread evenly on a ø6 mm petri-dish with Mueller-Hinton (MH) agar using sterile beads, and then 5 µg of methicillin sodium salt (Sigma, France) were dropped in the centre of the plate. Following the incubation at 37°C for 24 h, the diameter of zone of inhibition was measured.

## 3. Results and discussion

### 3.1. Design and validation of LAMP primers

Primers in the LAMP assay were designed to target a highly prevalent *mecA* gene among MRSA[25]. Five primer sets (Table S1) were screened for their specificity by conducting BLAST searches (Fig. S1), that indicated their high specificity to the *mecA* target. The primer sets were also validated for self-dimerization and hetero-dimer formation by using OligoAnalyzer. I*n silico* alignment with *mecA* sequence from non-*aureus Staphylococci* validated a high conservation rate of the gene among *Staphylococcus* spp. (Fig. S3). To select the best primer set we used gDNA of the MRSA strain JE2 and of the methicillin-sensitive *S. aureus* strain RN-R as a negative control. The amplification was recorded in a plate using a 10 ng/µL of gDNA and a fluorescent reporter. Typical sigmoidal amplification curves were observed in the reaction wells and result showed that the primer sets ID4 and ID16 accelerated the LAMP assay compared to other primers (Fig. 1a). The primer sets ID4 and ID16 enabled perceptible result for JE2 after a 25- to 30-min, meanwhile no effective amplification was detected at this time, using gDNA of RN-R (Fig. 1b).

**Fig. 1.**
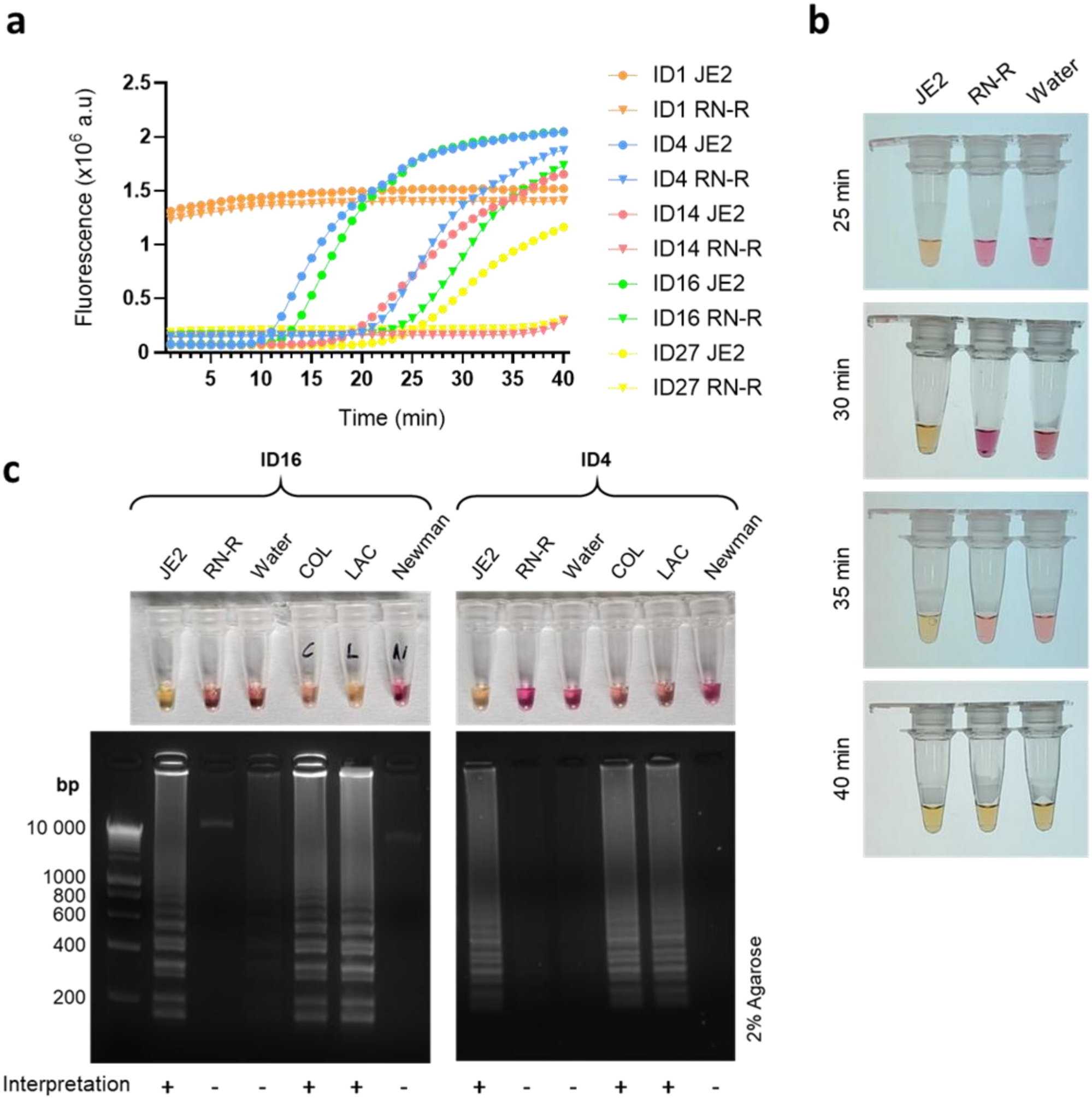
Validation of the LAMP primer sets. (a) The fluorescent LAMP assay performed using different primer sets with JE2 MRSA as positive control (●) and RN-R MSSA as a negative control (▼). (b) Colorimetric LAMP ID16 assay with JE2, RN-R and NTC at 25, 30, 35 and 40 min reaction time. (c) Colorimetric LAMP assay and 2% agarose gel electrophoresis of LAMP products obtained using primer sets ID4 and ID16 after 25 min, on MRSA strains (JE2, COL and LAC), MSSA strains (RN-R and Newman), and ultra-pure water as a no template control. SmartLadder DNA marker is shown on the left of the gel images.

Amplification of the negative controls have been observed with a delay in fluorometric LAMP. To validate 25-min reaction time, the amplification has been performed with colorimetric LAMP assay using MRSA JE2, MSSA RN-R and no template control for 25 min, 30 min, 35 min and 40 min (Fig. S4). Only the tube with JE2 turned to yellow at 25 and 30 min, while negatives RN-R and NTC slightly turned to yellow at 40 min, confirming the 25-min reaction time. The weak late amplification, that sometimes occurs in LAMP assays, reflects an intrinsic property of LAMP reactions, which can produce nonspecific signals if the reaction extends beyond the optimized time [16, 26-28].

Further verification of the specificity of the primer sets ID4 and ID16, included the cross reactivity between well-characterized *S. aureus* strains holding *mecA* gene and sensitive *S. aureus* strains (Fig. 1b). The same concentration of gDNA (10 ng/µL) for each bacterial strain was used to evaluate the colorimetric read-out. The reaction mixtures changed colour from pink to yellow within 25 min, indicating the amplification, while no colour change was observed with negative controls (Fig. 1b). When the reaction mixtures were loaded onto the gel, their electrophoretic bands confirmed the specificity of the primer sets ID4 and ID16 (Fig. 1b). Interestingly, a clear amplification was observed with *S. aureus* LAC strain that holds one mismatch in *mecA* sequence (position 1271, C to T) (Fig. S3).

To verify the optimal amplification temperature of the LAMP assay, we used gDNA from JE2 MRSA (10 ng/µL) and the primer set ID16, to perform reactions at various temperatures (63–66 °C, with 1°C intervals). Amplification was observed at all tested temperatures (Fig. S5). Ultimately, 65 °C, the temperature recommended by the LAMP kit supplier, was selected as the optimal amplification temperature, and was further used.

### 3.2. Limit of detection of the LAMP assay

To determinate the limit of detection (LoD) of the established LAMP assay, serial 10-fold dilutions (ranging from 1.5 pg to 1.5 fg of gDNA/µL) were tested using the primer sets ID4 and ID 16 (Fig. 2). Colorimetric assay indicated LoD of 1.5 pg gDNA/µL (corresponding to *ca*. 500 copies of *mecA*/µL) and 15 fg gDNA/µL (*ca*. 5 copies of *mecA*/µL) using the primer sets ID4 and ID16, respectively (Fig. 2a, b). These LoD values were confirmed when the LAMP assay was coupled with the fluorescent read-out (Fig. 2c, d). Since the lower LoD was obtained with the primer set ID16 (*ca*. 5 copies of *mecA*/µL), this set was further use. Since there is one copy of *mecA* gene in the *S. aureus* genome, the calculated LoD of the assay is 5×10^3^ CFU/mL. For comparison, when the amplification was performed by qPCR, using JE2 strain, the LOD of 1 pg/µL of gDNA (corresponding to 340 *mecA* copies/µL) was obtained (Fig. S6). The qPCR standard curve had a good linear correlation between the DNA concentration and the C_t_ value (R^2^ = 0.995), indicating that the developed qPCR assay was reliable and could quantify the target sequence. However, the LAMP assay was more rapid, easier to perform and lower in cost. Therefore, LAMP is more suitable for quantitative analysis of samples with low target concentrations. Furthermore, the low LoD of our LAMP assay is comparable with values obtained for *mecA* detection using other LAMP assay as listed in Table 1, recently proposed in the literature. To further evaluate the developed colorimetric LAMP assay, we applied it on food and clinical *S. aureus* isolates and compared the results with the qPCR, as illustrated in Fig. 3a.

**Table 1.** Performance comparison of the developed LAMP assay with previously reported LAMP assays for *mecA* detection, reported in literature.

| **Detection method** | **LoD (CFU/mL)** | **LoD (pg/µL)** | **Detection time (min)** | **Reference** |
|---|---|---|---|---|
| LAMP | NR | NR | 60 | <sup>29</sup> |
| LAMP | NR | NR | 60 | <sup>30</sup> |
| PCR | NR | NR | NR | <sup>30</sup> |
| LAMP | NR | NR | 60 | <sup>31</sup> |
| LAMP | 1.5×10<sup>7</sup> | 4.6 | 40 | <sup>32</sup> |
| LAMP | 100 | NR | 60 | <sup>33</sup> |
| PCR | 1x10<sup>3</sup> | NR | NR | <sup>33</sup> |
| LAMP | NR | NR | 50-60 | <sup>34</sup> |
| LAMP | NR | NR | 60 | <sup>35</sup> |
| LAMP | NR | NR | 65 | <sup>36</sup> |
| LAMP | NR | NR | 30-40 | <sup>37</sup> |
| LAMP | 1.5×10<sup>4</sup> | NR | 65 | <sup>38</sup> |
| LAMP | 10<sup>5</sup> | NR | 60 | <sup>39</sup> |
| LAMP | 5x10<sup>3</sup> | 0.015 | 25 | Our work |
| qPCR | 3.4x10<sup>6</sup> | 1 | 60 | Our work |

NR, not reported by authors

**Fig. 2.** Sensitivity of the LAMP assay with the primer set ID4 and ID16 tested using gDNA of JE2 strain carrying the *mecA* gene and colorimetric (a, b) and fluorometric (c, d) read out using indicated amounts of gDNA/reaction. gDNA of methicillin sensitive RN-R strain (200 ng) was used as a negative control. The bars represent the standard deviation of a quadruplet.

**Fig. 3.**
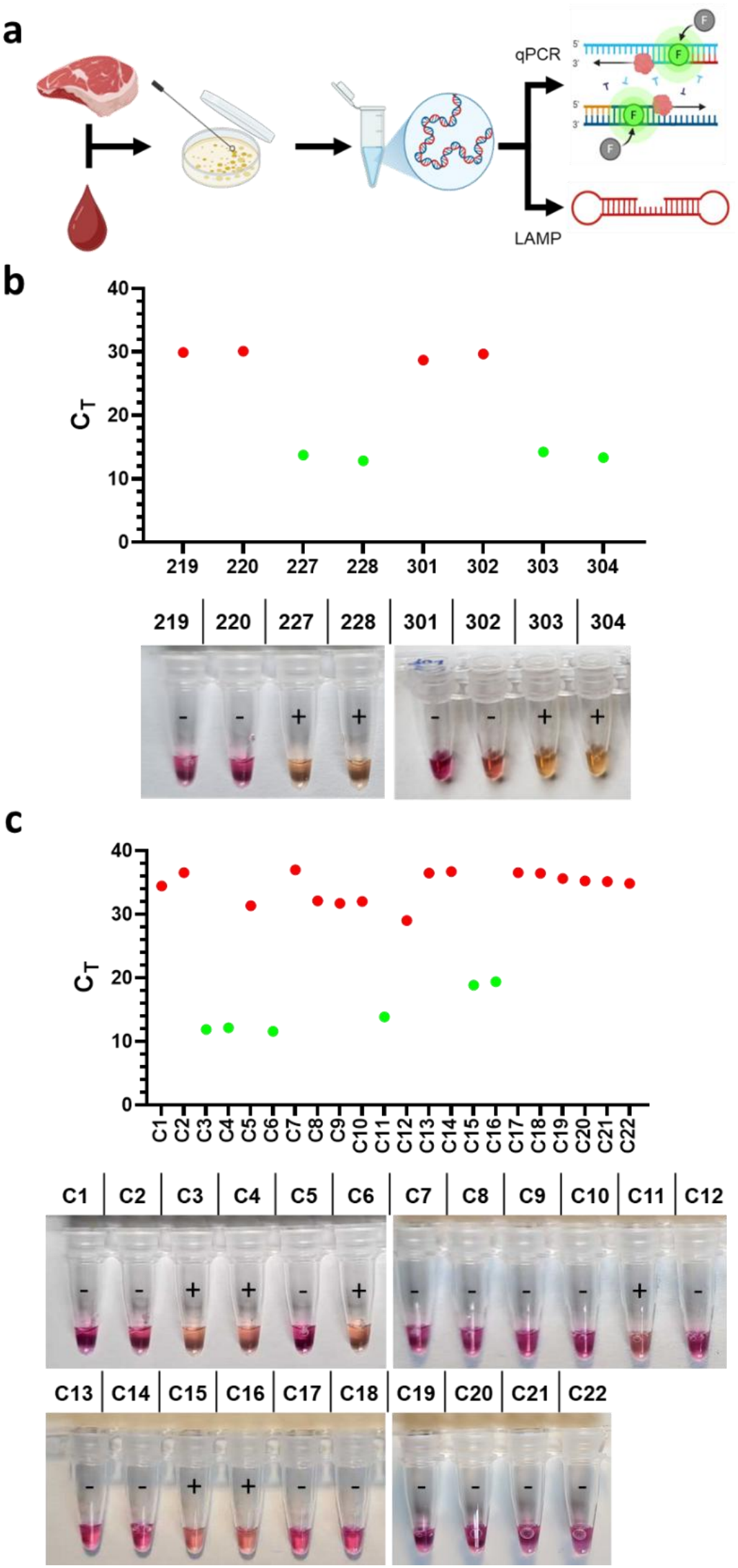
Comparison of qPCR and LAMP assay for detection of the *mecA* gene. (a) Illustration of the experimental approach. Food and blood *S. aureus* isolates were confirmed and their gDNA after extraction were tested in parallel by qPCR and LAMP with the ID16 primer set. (b) Results obtained using the food isolates. (c) Results obtained using the clinical blood isolates.

### 3.3. LAMP assay on food samples

Eight *S. aureus* strains isolated from raw meat and milk samples were first tested for their susceptibility to methicillin. Overnight bacterial solutions were diluted and plated on MH solid medium, and methicillin (5 µg) was spotted in the center of plates, which were incubated and photographed. Four strains showed resistance (milk isolates 227, 228, and meat isolates 303, 304), while others (milk isolates 219, 220, and meat isolates 301, 302) were susceptible and showed well-defined zones of inhibition (Fig. S7). To verify whether the observed β-lactam resistance was *mecA*-mediated, the qPCR analysis was performed. As shown in Fig. 3b, all resistant samples showed the amplification of *mecA* gene. In the LAMP assay, 4 food samples testing positive by the two conventional microbiological methods were also positive as colour of their solution changed from pink to yellow (Fig. 3b).

When methicillin susceptibility test was performed on *S. aureus* blood isolates, 6 out of 22 strains had well defined inhibition zone, without apparition of resistant colonies inside the zone, and were identified as MRSA (Fig. S8). qPCR confirmed that all 6 resistant strains possess the *mecA* gene (Fig. 3c). A pink-to-yellow colour change was observed with the same 6 out of 22 isolates when tested using the developed LAMP assay (Fig. 3c). Results of the colorimetric LAMP assays were confirmed by running the reaction products on a 2% agarose gel (Fig. S9).

Overall, our LAMP assay correctly identified both *mecA*-positive and *mecA*-negative samples in food and clinical isolates, with 100% specificity (correct positive and negative results relative to the total number of tests performed), as confirmed by the reference method, qPCR (Table S3).

### 3.4 Direct LAMP assay in serum

Bovine serum samples spiked with JE2 or RN-R strains was analysed under optimized conditions to evaluate the matrix effect on overall analytical performance (Fig. 4). Both colorimetric and fluorescent detections clearly distinguished between *mecA*-positive and *mecA*-negative strains when performed directly on bacterial lysates without DNA purification, as shown in Figs. 4b and 4c. This finding demonstrating the robustness of the developed method.

**Fig. 4.**
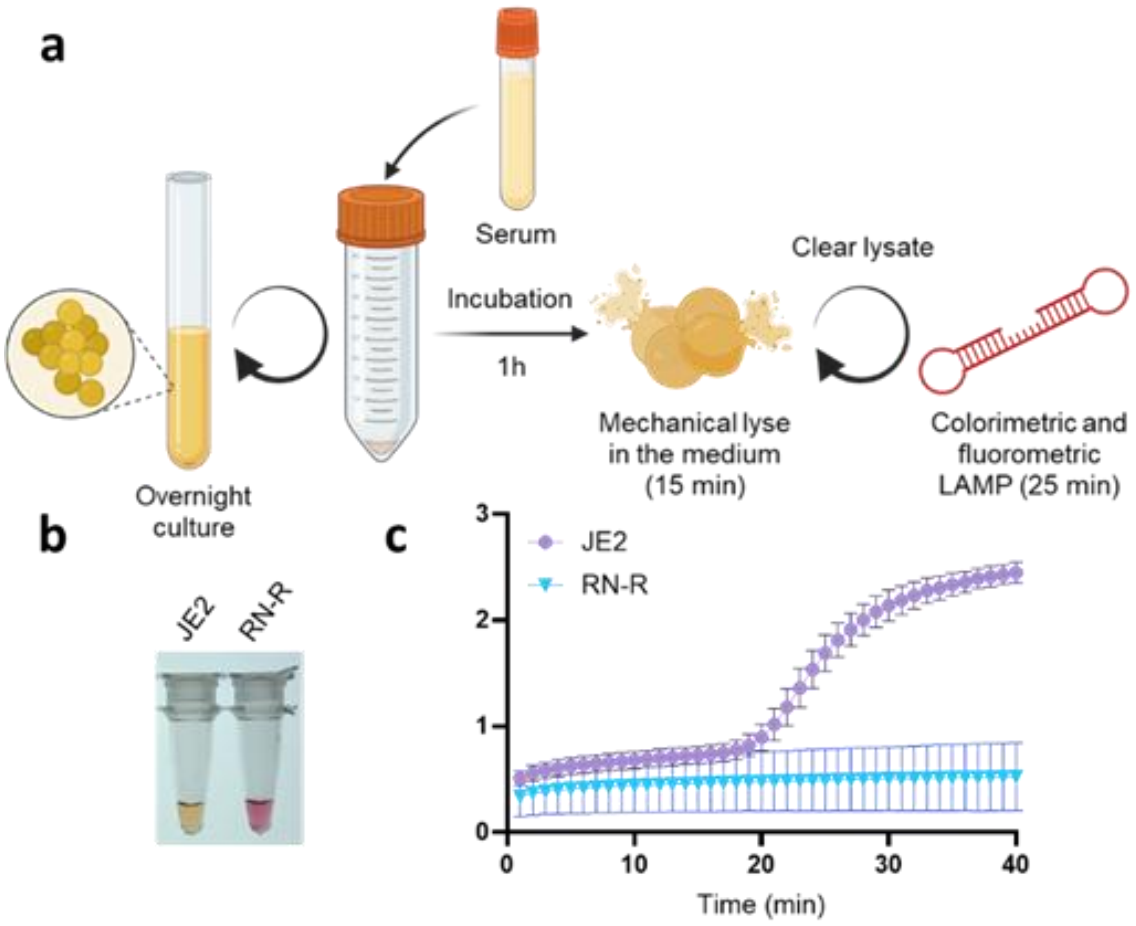
Spiked serum analytical sensitivity on LAMP *mecA*-positive (JE2) and *mecA*-negative (RN-R) strains. (a) Procedure of serum spiking and its analysis. Spiked serum analysed using (b) colorimetric and (c) fluorometric LAMP assay.

## 4. Conclusions

We successfully developed a LAMP assay for the detection of the *mecA* gene in methicillin-resistant *S. aureus* strains. The specificity of newly designed LAMP primers was confirmed *in silico* against *mecA* gene sequences from other non-*S. aureus* staphylococci. The colorimetric and fluorometric LAMP assay enabled reliable identification of *mecA*-positive genomic DNA samples through a simple procedure: direct mixing of the target DNA with the reaction solution and incubation at 65 °C for 25 minutes. In addition to its rapid turnaround time, the assay exhibited excellent performance in terms of limit of detection (5 copies/µL) and specificity (100%). These characteristics directly address the limitations of conventional PCR-based methods and highlight the potential of LAMP for use in point-of-care settings, where rapid and accessible diagnostics are essential. Using both food and clinical *S. aureus* isolates, we demonstrated that the specificity of the LAMP assay was comparable to qPCR. However, unlike qPCR, the LAMP assay was significantly easier to perform and more cost-effective. Furthermore, the robustness of the assay was validated in complex matrices serum and milk, where direct mechanical lysis was used and no significant inhibition was observed in colorimetric assay. The developed LAMP assay, thus, offers a scalable and cost-effective solution for decentralized diagnostics. Taken together, these findings show that the developed LAMP assay provides a practical, scalable, and affordable solution for decentralized and point-of-care diagnostics in a timely manner.

## Data Availability

All data produced in the present work are contained in the manuscript

## Author contributions

L.B.: Methodology, Investigation, Writing - original draft, Writing - review & editing. M.D.: Methodology, Investigation, Writing - original draft, Writing - review & editing. D.P. Writing - original draft, Writing - review & editing. M.M.: Writing - review & editing. I.G.: Funding acquisition, Writing - review & editing. J.V. Conceptualization, Funding acquisition, Project administration. Writing - original draft, Writing - review & editing.

## Conflicts of interest

There are no conflicts to declare.

## Data availability

The data supporting this article have been included as part of the Electronic Supplementary Information.

## Fundings

This work is supported by the European Union under grant agreement N° 101135402 (MOBILES project), and under grant agreement N° 872662 (IPANEMA project).

## Notes

### Competing Interest Statement

The authors have declared no competing interest.

