## Supplementary Information for "A rapid and sensitive Loop-Mediated Isothermal Amplification (LAMP) assay for detection of *mecA* antibiotic resistance gene in clinical and food bacterial isolates"

**Summary**

Table S1. LAMP primer sets

Table S2. Accession number of *Staphylococcus spp.* used for *in silico* alignment

Table S3. Comparison of the *mecA*-screening on food and blood isolates

Figure S1. Workflow showing the experimental plan

Figure S2. *In silico* hybridization of LAMP primer set ID16 within *mecA* sequence

Figure S3. Alignment of ID4 and ID16 on *S. aureus* JE2 compared with other *mecA* sequences

Figure S4. Determination of the reaction time

Figure S5. Fluorescent LAMP reaction at different temperatures

Figure S6. Sensitivity of the qPCR assay

Figure S7. Methicillin susceptibility test on *S. aureus* food isolates

Figure S8. Methicillin susceptibility test on *S. aureus* clinical blood isolates

Figure S9. Validation of the colorimetric LAMP by agarose gel electrophoresis

Table S1. LAMP primers used in this study.

| **Name** | **Sequence** (5'→3') |
|---|---|
| ID1-F3 | CCTTTACCATCGATTTTATAACTTG |
| ID1-B3 | GCACACCTTCATATGACGT |
| ID1-FIP (F1c + F2) | CAGATTACAACTTCACCAGGTTCAA-TTGTTATTTAACCCAATCATTGC |
| ID1-BIP (B1c + B2) | GAACTTGTTGAGCAGAGGTTCT-CTATCCATTTATGTATGGCATGAG |
| ID4-F3 | TTAACCCAATCATTGCTGTT |
| ID4-B3 | GCTATCCACCCTCAAACAG |
| ID4-FIP (F1c + F2) | CCGAAGATAAAAAAGAACCTCTGCT-AATATTTTTTGAGTTGAACCTGG |
| ID4-BIP (B1c + B2) | ATTCTTCGTTACTCATGCCATACA-AGCACTTGTAAGCACACC |
| ID14-F3 | GCCATCTTTTTTCTTTTTCTCT |
| ID14-B3 | CCTCTAGGAAAAGCGACTTC |
| ID14-FIP (F1c + F2) | AGCTCCAACATGAAGATGGCT-GCGATTGTATTGCTATTATCGTC |
| ID14-BIP (B1c + B2) | TTCGAGTCCCTTTTTACCAATAACT-TGGTCCCATTAACTCTGAAG |
| ID16-F3 | CGTCAACGATTGTGACAC |
| ID16-B3 | CAACTAATGAAACAGAAAGTCG |
| ID16-FIP (F1c + F2) | GCAGTTATTGGTAAAAAGGGACTCG-GATAGCCATCTTCATGTTGGA |
| ID16 -BIP (B1c + B2) | AATTCTTCAGAGTTAATGGGACCAA-TATCCTCTAGGAAAAGCGACT |
| ID27-F3 | GTAAAGTTTTTCGAGTCCCTT |
| ID27-B3 | GGATGAATATTTAAGTGATTTCGCA |
| ID27-FIP (F1c + F2) | TTGGTCCCATTAACTCTGAAGAATT-ATAACTGCATCATCTTTATAGCCT |
| ID27-BIP (B1c + B2) | AACCTAATAGATGTGAAGTCGCTTT-CTTACAACTAATGAAACAGAAAGTC |

Table S2. Accession number of *Staphylococcus* spp. used for the *in silico* alignment

| ***S. argenteus*** | | ***S. aureus*** | | ***S. capitis*** | | ***S. caprae*** | |
|---|---|---|---|---|---|---|---|
| **Strain** | **Accession** | **Strain** | **Accession** | **Strain** | **Accession** | **Strain** | **Accession** |
| / | NC_016941.1 | JE2 | NZ_CP020619.1 | XWZ | CP098820.1 | JMUB898 | NZ_AP018587.1 |
| *RIVM_M020832* | CP086570.1 | COL | NC_002951.2 | CSLA5 | NG_047941.1 | *JMUB590* | AP018586.1 |
| *PHL3432* | PUXC01000007.1 | LAC | CP055225.1 | urealyticus | CP145228.1 | | |
| *CCUG* | NSBX01000054.1 | BSN124 | CP149937.1 | LR 95 | CP092857.1 | | |
| *MRSA-139-14-168* | CP096114.1 | Sau1 | CP141522.1 | LZD8 | CP098816.1 | | |

| ***S. epidermidis*** | | ***S. haemolyticus*** | | ***S. hominis*** | | ***S. lugdunensis*** | |
|---|---|---|---|---|---|---|---|
| **Strain** | **Accession** | **Strain** | **Accession** | **Strain** | **Accession** | **Strain** | **Accession** |
| 6593473 | CP196900.1 | GDY8P80P | CP063443.1 | C5 | CP093539.1 | RMLUG2 | CP084434.1 |
| pb1 | PV890966.1 | VB5326 | CP045137.2 | acrlw | CP134693.1 | RMLUG1 | CP084480.1 |
| B1276912 | CP073865.1 | VB19458 | CP045187.2 | Sh8213 | CP142855.1 | RMLUG6 | CP084444.1 |
| UMCG350 | CP064603.1 | FDAARGOS_517 | CP033814.1 | FDAARGOS_661 | CP054550.1 | SO-SLU20-1 | CP102431.1 |
| HD21N4 | CP060157.1 | SGAir0252 | CP025031.1 | acrlk | CP090014.1 | RMLUG3 | CP084437.1 |
| AR4072 | CP133663.1 | | | | | | |

Table S3. Comparison of the *mecA*-screening on food and blood isolates.

| LAMP assay | | qPCR assay: Positive | qPCR assay: Negative | qPCR assay: Total |
|---|---|---|---|---|
| | **Positive** | 10 | 0 | 10 |
| | **Negative** | 0 | 20 | 20 |
| | **Total** | 10 | 20 | **30** |

Accuracy: (10+20)/30= 100%
Sensitivity: 10/10=100%
Specificity: 20/20=100%

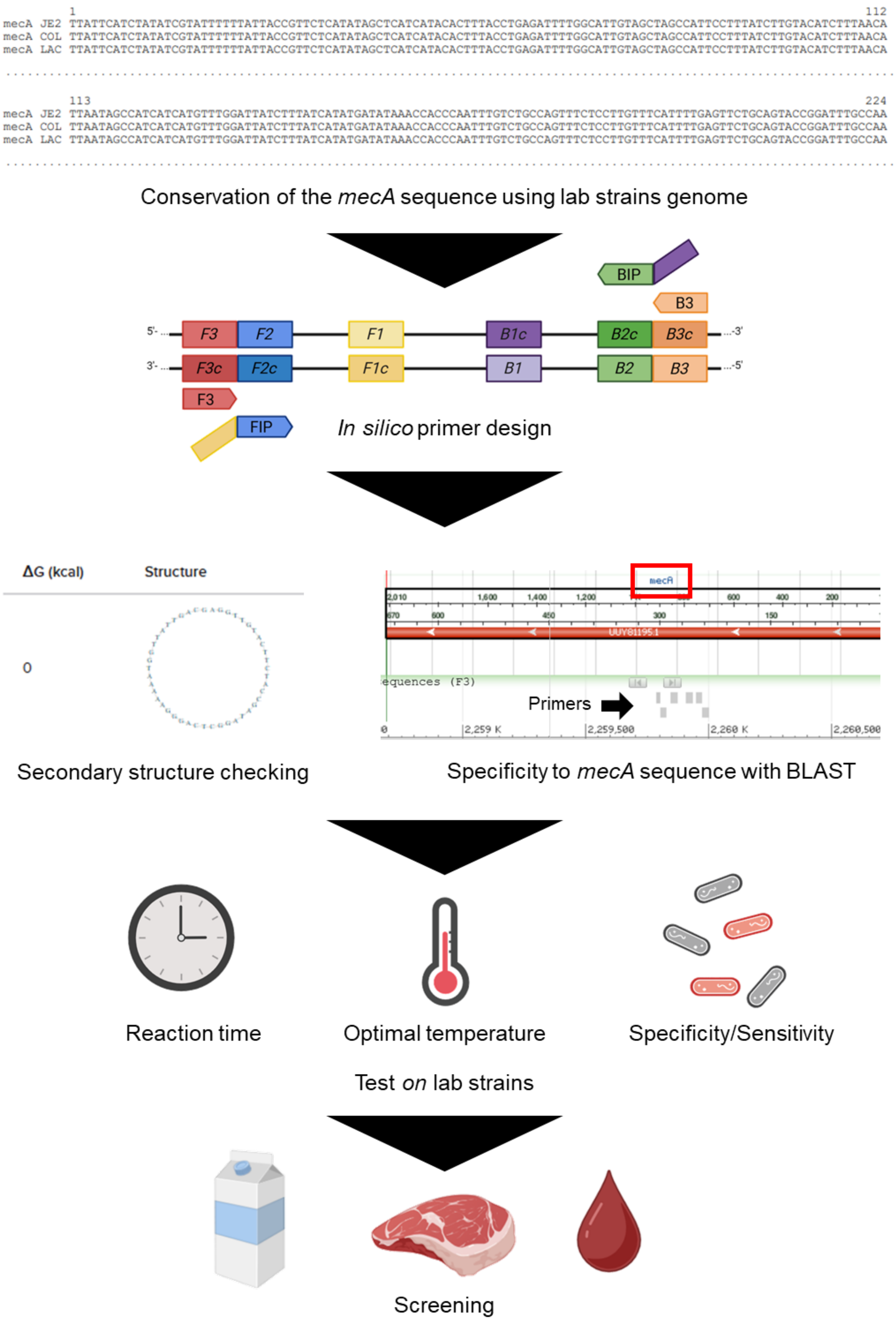


Figure S1. Workflow showing the experimental step-by-step plan for developing an efficient LAMP assay for *mecA* gene detection.

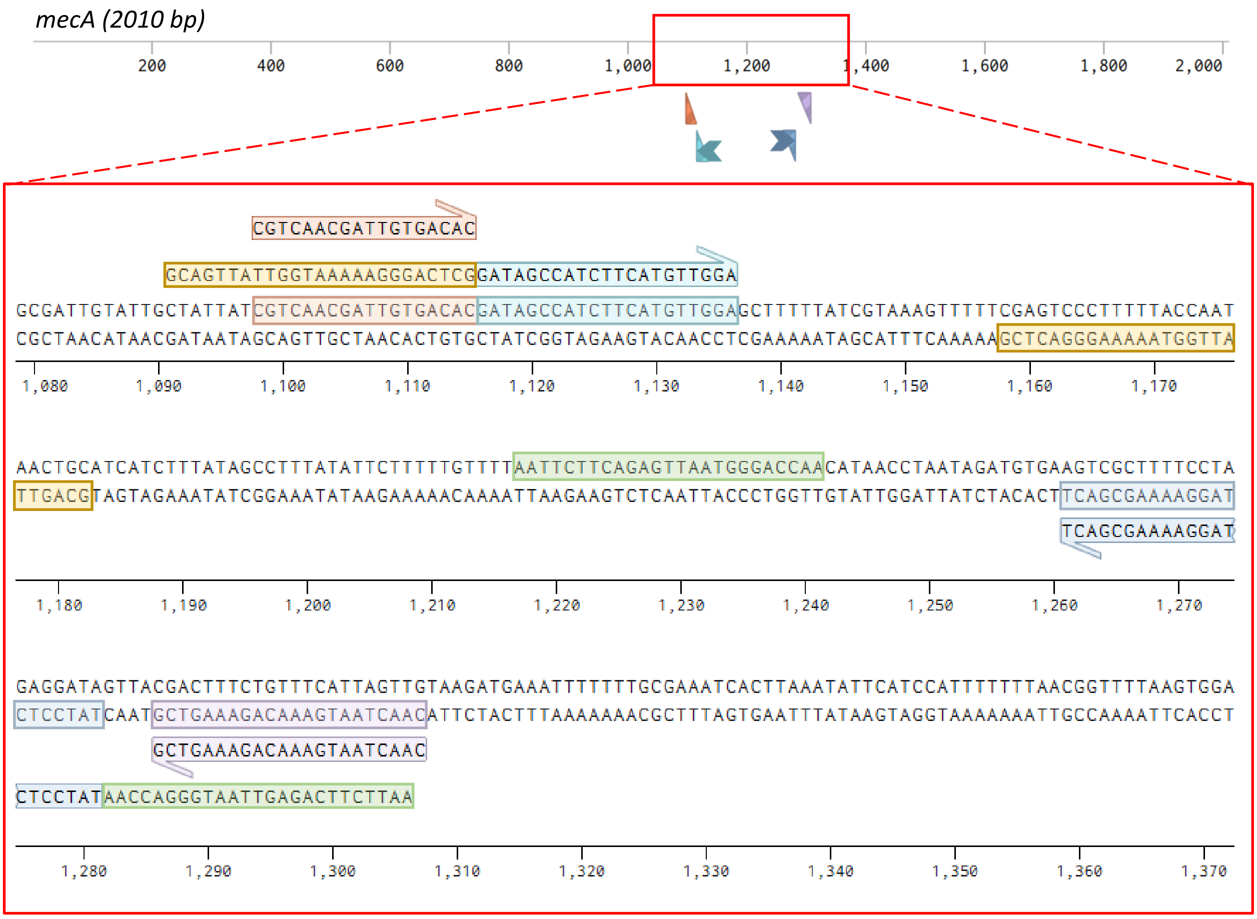


Figure S2. *In silico* hybridization of LAMP primer set ID16 positioned to *mecA* sequence (2010 bp, GenBank accession number NZ_CP020619): F3 (1098–1115 bp), F2 (1116–1136 bp), F1c (1158–1182 bp), B1c (1217–1241 bp), B2 (1261–1281 bp) and B3 (1286–1307 bp).

**a**

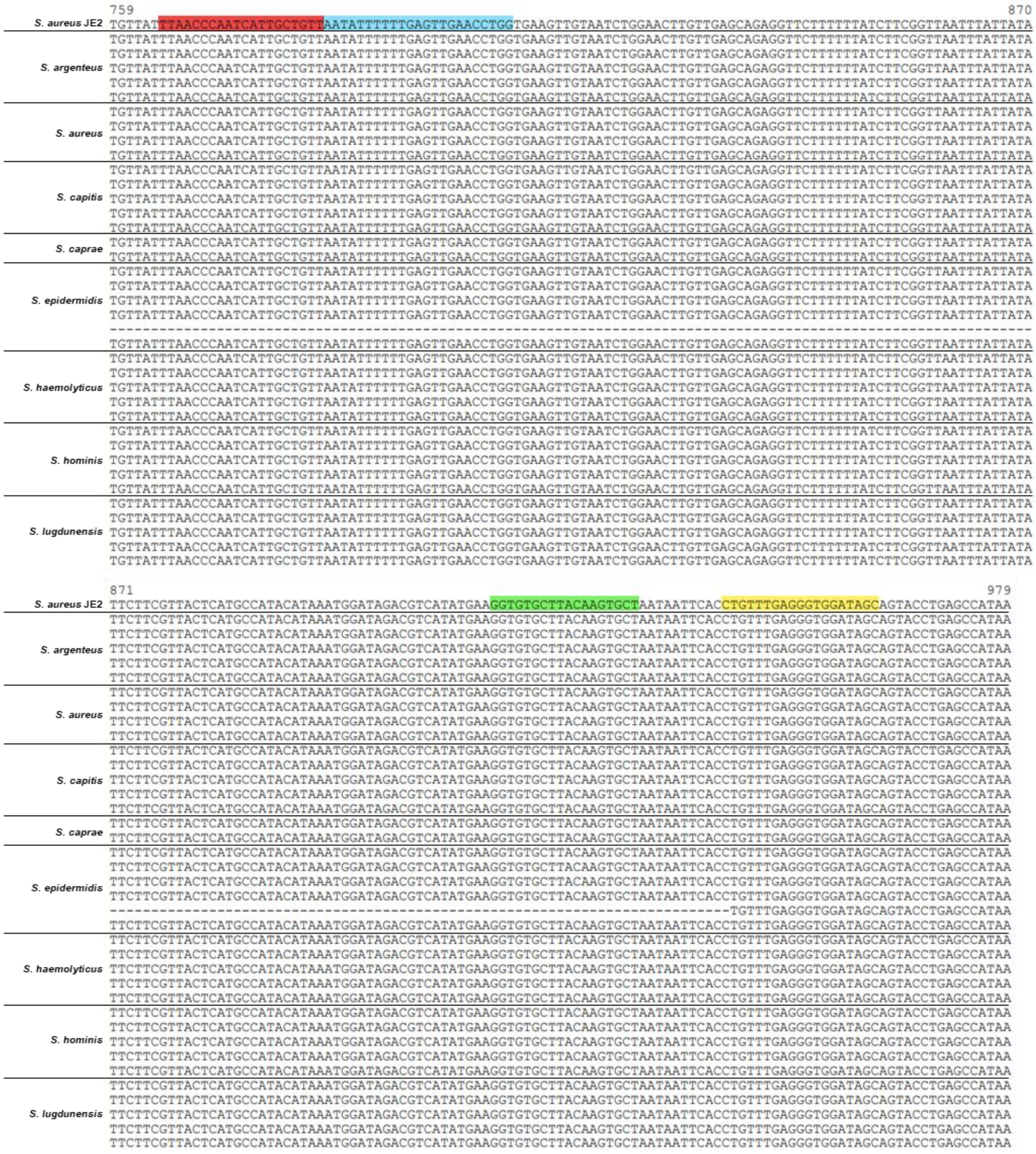

759
870
S. aureus JE2 TGTTATTTAACCCAATCATTGCTGTTAATATTTTTTGAGTTGAACCTGGTGAAGTTGTAATCTGGAACTTGTTGAGCAGAGGTTCTTTTTTATCTTCGGTTAATTTATTATA
S. argenteus
TGTTATTTAACCCAATCATTGCTGTTAATATTTTTTGAGTTGAACCTGGTGAAGTTGTAATCTGGAACTTGTTGAGCAGAGGTTCTTTTTTATCTTCGGTTAATTTATTATA
TGTTATTTAACCCAATCATTGCTGTTAATATTTTTTGAGTTGAACCTGGTGAAGTTGTAATCTGGAACTTGTTGAGCAGAGGTTCTTTTTTATCTTCGGTTAATTTATTATA
TGTTATTTAACCCAATCATTGCTGTTAATATTTTTTGAGTTGAACCTGGTGAAGTTGTAATCTGGAACTTGTTGAGCAGAGGTTCTTTTTTATCTTCGGTTAATTTATTATA
TGTTATTTAACCCAATCATTGCTGTTAATATTTTTTGAGTTGAACCTGGTGAAGTTGTAATCTGGAACTTGTTGAGCAGAGGTTCTTTTTTATCTTCGGTTAATTTATTATA
TGTTATTTAACCCAATCATTGCTGTTAATATTTTTTGAGTTGAACCTGGTGAAGTTGTAATCTGGAACTTGTTGAGCAGAGGTTCTTTTTTATCTTCGGTTAATTTATTATA
S. aureus
TGTTATTTAACCCAATCATTGCTGTTAATATTTTTTGAGTTGAACCTGGTGAAGTTGTAATCTGGAACTTGTTGAGCAGAGGTTCTTTTTTATCTTCGGTTAATTTATTATA
TGTTATTTAACCCAATCATTGCTGTTAATATTTTTTGAGTTGAACCTGGTGAAGTTGTAATCTGGAACTTGTTGAGCAGAGGTTCTTTTTTATCTTCGGTTAATTTATTATA
TGTTATTTAACCCAATCATTGCTGTTAATATTTTTTGAGTTGAACCTGGTGAAGTTGTAATCTGGAACTTGTTGAGCAGAGGTTCTTTTTTATCTTCGGTTAATTTATTATA
TGTTATTTAACCCAATCATTGCTGTTAATATTTTTTGAGTTGAACCTGGTGAAGTTGTAATCTGGAACTTGTTGAGCAGAGGTTCTTTTTTATCTTCGGTTAATTTATTATA
S. capitis
TGTTATTTAACCCAATCATTGCTGTTAATATTTTTTGAGTTGAACCTGGTGAAGTTGTAATCTGGAACTTGTTGAGCAGAGGTTCTTTTTTATCTTCGGTTAATTTATTATA
TGTTATTTAACCCAATCATTGCTGTTAATATTTTTTGAGTTGAACCTGGTGAAGTTGTAATCTGGAACTTGTTGAGCAGAGGTTCTTTTTTATCTTCGGTTAATTTATTATA
TGTTATTTAACCCAATCATTGCTGTTAATATTTTTTGAGTTGAACCTGGTGAAGTTGTAATCTGGAACTTGTTGAGCAGAGGTTCTTTTTTATCTTCGGTTAATTTATTATA
TGTTATTTAACCCAATCATTGCTGTTAATATTTTTTGAGTTGAACCTGGTGAAGTTGTAATCTGGAACTTGTTGAGCAGAGGTTCTTTTTTATCTTCGGTTAATTTATTATA
TGTTATTTAACCCAATCATTGCTGTTAATATTTTTTGAGTTGAACCTGGTGAAGTTGTAATCTGGAACTTGTTGAGCAGAGGTTCTTTTTTATCTTCGGTTAATTTATTATA
S. caprae
TGTTATTTAACCCAATCATTGCTGTTAATATTTTTTGAGTTGAACCTGGTGAAGTTGTAATCTGGAACTTGTTGAGCAGAGGTTCTTTTTTATCTTCGGTTAATTTATTATA
TGTTATTTAACCCAATCATTGCTGTTAATATTTTTTGAGTTGAACCTGGTGAAGTTGTAATCTGGAACTTGTTGAGCAGAGGTTCTTTTTTATCTTCGGTTAATTTATTATA
S. epidermidis
TGTTATTTAACCCAATCATTGCTGTTAATATTTTTTGAGTTGAACCTGGTGAAGTTGTAATCTGGAACTTGTTGAGCAGAGGTTCTTTTTTATCTTCGGTTAATTTATTATA
TGTTATTTAACCCAATCATTGCTGTTAATATTTTTTGAGTTGAACCTGGTGAAGTTGTAATCTGGAACTTGTTGAGCAGAGGTTCTTTTTTATCTTCGGTTAATTTATTATA
TGTTATTTAACCCAATCATTGCTGTTAATATTTTTTGAGTTGAACCTGGTGAAGTTGTAATCTGGAACTTGTTGAGCAGAGGTTCTTTTTTATCTTCGGTTAATTTATTATA
TGTTATTTAACCCAATCATTGCTGTTAATATTTTTTGAGTTGAACCTGGTGAAGTTGTAATCTGGAACTTGTTGAGCAGAGGTTCTTTTTTATCTTCGGTTAATTTATTATA
--------------------------------------------------------------------------------------------------------------
TGTTATTTAACCCAATCATTGCTGTTAATATTTTTTGAGTTGAACCTGGTGAAGTTGTAATCTGGAACTTGTTGAGCAGAGGTTCTTTTTTATCTTCGGTTAATTTATTATA
S. haemolyticus
TGTTATTTAACCCAATCATTGCTGTTAATATTTTTTGAGTTGAACCTGGTGAAGTTGTAATCTGGAACTTGTTGAGCAGAGGTTCTTTTTTATCTTCGGTTAATTTATTATA
TGTTATTTAACCCAATCATTGCTGTTAATATTTTTTGAGTTGAACCTGGTGAAGTTGTAATCTGGAACTTGTTGAGCAGAGGTTCTTTTTTATCTTCGGTTAATTTATTATA
TGTTATTTAACCCAATCATTGCTGTTAATATTTTTTGAGTTGAACCTGGTGAAGTTGTAATCTGGAACTTGTTGAGCAGAGGTTCTTTTTTATCTTCGGTTAATTTATTATA
TGTTATTTAACCCAATCATTGCTGTTAATATTTTTTGAGTTGAACCTGGTGAAGTTGTAATCTGGAACTTGTTGAGCAGAGGTTCTTTTTTATCTTCGGTTAATTTATTATA
TGTTATTTAACCCAATCATTGCTGTTAATATTTTTTGAGTTGAACCTGGTGAAGTTGTAATCTGGAACTTGTTGAGCAGAGGTTCTTTTTTATCTTCGGTTAATTTATTATA
S. hominis
TGTTATTTAACCCAATCATTGCTGTTAATATTTTTTGAGTTGAACCTGGTGAAGTTGTAATCTGGAACTTGTTGAGCAGAGGTTCTTTTTTATCTTCGGTTAATTTATTATA
TGTTATTTAACCCAATCATTGCTGTTAATATTTTTTGAGTTGAACCTGGTGAAGTTGTAATCTGGAACTTGTTGAGCAGAGGTTCTTTTTTATCTTCGGTTAATTTATTATA
TGTTATTTAACCCAATCATTGCTGTTAATATTTTTTGAGTTGAACCTGGTGAAGTTGTAATCTGGAACTTGTTGAGCAGAGGTTCTTTTTTATCTTCGGTTAATTTATTATA
TGTTATTTAACCCAATCATTGCTGTTAATATTTTTTGAGTTGAACCTGGTGAAGTTGTAATCTGGAACTTGTTGAGCAGAGGTTCTTTTTTATCTTCGGTTAATTTATTATA
TGTTATTTAACCCAATCATTGCTGTTAATATTTTTTGAGTTGAACCTGGTGAAGTTGTAATCTGGAACTTGTTGAGCAGAGGTTCTTTTTTATCTTCGGTTAATTTATTATA
S. lugdunensis
TGTTATTTAACCCAATCATTGCTGTTAATATTTTTTGAGTTGAACCTGGTGAAGTTGTAATCTGGAACTTGTTGAGCAGAGGTTCTTTTTTATCTTCGGTTAATTTATTATA
TGTTATTTAACCCAATCATTGCTGTTAATATTTTTTGAGTTGAACCTGGTGAAGTTGTAATCTGGAACTTGTTGAGCAGAGGTTCTTTTTTATCTTCGGTTAATTTATTATA
TGTTATTTAACCCAATCATTGCTGTTAATATTTTTTGAGTTGAACCTGGTGAAGTTGTAATCTGGAACTTGTTGAGCAGAGGTTCTTTTTTATCTTCGGTTAATTTATTATA
TGTTATTTAACCCAATCATTGCTGTTAATATTTTTTGAGTTGAACCTGGTGAAGTTGTAATCTGGAACTTGTTGAGCAGAGGTTCTTTTTTATCTTCGGTTAATTTATTATA
TGTTATTTAACCCAATCATTGCTGTTAATATTTTTTGAGTTGAACCTGGTGAAGTTGTAATCTGGAACTTGTTGAGCAGAGGTTCTTTTTTATCTTCGGTTAATTTATTATA
871
979
S. aureus JE2 TTCTTCGTTACTCATGCCATACATAAATGGATAGACGTCATATGAAGGTGTGCTTACAAGTGCTAATAATTCACCTGTTTGAGGGTGGATAGCAGTACCTGAGCCATAA
S. argenteus
TTCTTCGTTACTCATGCCATACATAAATGGATAGACGTCATATGAAGGTGTGCTTACAAGTGCTAATAATTCACCTGTTTGAGGGTGGATAGCAGTACCTGAGCCATAA
TTCTTCGTTACTCATGCCATACATAAATGGATAGACGTCATATGAAGGTGTGCTTACAAGTGCTAATAATTCACCTGTTTGAGGGTGGATAGCAGTACCTGAGCCATAA
TTCTTCGTTACTCATGCCATACATAAATGGATAGACGTCATATGAAGGTGTGCTTACAAGTGCTAATAATTCACCTGTTTGAGGGTGGATAGCAGTACCTGAGCCATAA
TTCTTCGTTACTCATGCCATACATAAATGGATAGACGTCATATGAAGGTGTGCTTACAAGTGCTAATAATTCACCTGTTTGAGGGTGGATAGCAGTACCTGAGCCATAA
TTCTTCGTTACTCATGCCATACATAAATGGATAGACGTCATATGAAGGTGTGCTTACAAGTGCTAATAATTCACCTGTTTGAGGGTGGATAGCAGTACCTGAGCCATAA
S. aureus
TTCTTCGTTACTCATGCCATACATAAATGGATAGACGTCATATGAAGGTGTGCTTACAAGTGCTAATAATTCACCTGTTTGAGGGTGGATAGCAGTACCTGAGCCATAA
TTCTTCGTTACTCATGCCATACATAAATGGATAGACGTCATATGAAGGTGTGCTTACAAGTGCTAATAATTCACCTGTTTGAGGGTGGATAGCAGTACCTGAGCCATAA
TTCTTCGTTACTCATGCCATACATAAATGGATAGACGTCATATGAAGGTGTGCTTACAAGTGCTAATAATTCACCTGTTTGAGGGTGGATAGCAGTACCTGAGCCATAA
TTCTTCGTTACTCATGCCATACATAAATGGATAGACGTCATATGAAGGTGTGCTTACAAGTGCTAATAATTCACCTGTTTGAGGGTGGATAGCAGTACCTGAGCCATAA
S. capitis
TTCTTCGTTACTCATGCCATACATAAATGGATAGACGTCATATGAAGGTGTGCTTACAAGTGCTAATAATTCACCTGTTTGAGGGTGGATAGCAGTACCTGAGCCATAA
TTCTTCGTTACTCATGCCATACATAAATGGATAGACGTCATATGAAGGTGTGCTTACAAGTGCTAATAATTCACCTGTTTGAGGGTGGATAGCAGTACCTGAGCCATAA
TTCTTCGTTACTCATGCCATACATAAATGGATAGACGTCATATGAAGGTGTGCTTACAAGTGCTAATAATTCACCTGTTTGAGGGTGGATAGCAGTACCTGAGCCATAA
TTCTTCGTTACTCATGCCATACATAAATGGATAGACGTCATATGAAGGTGTGCTTACAAGTGCTAATAATTCACCTGTTTGAGGGTGGATAGCAGTACCTGAGCCATAA
TTCTTCGTTACTCATGCCATACATAAATGGATAGACGTCATATGAAGGTGTGCTTACAAGTGCTAATAATTCACCTGTTTGAGGGTGGATAGCAGTACCTGAGCCATAA
S. caprae
TTCTTCGTTACTCATGCCATACATAAATGGATAGACGTCATATGAAGGTGTGCTTACAAGTGCTAATAATTCACCTGTTTGAGGGTGGATAGCAGTACCTGAGCCATAA
TTCTTCGTTACTCATGCCATACATAAATGGATAGACGTCATATGAAGGTGTGCTTACAAGTGCTAATAATTCACCTGTTTGAGGGTGGATAGCAGTACCTGAGCCATAA
S. epidermidis
TTCTTCGTTACTCATGCCATACATAAATGGATAGACGTCATATGAAGGTGTGCTTACAAGTGCTAATAATTCACCTGTTTGAGGGTGGATAGCAGTACCTGAGCCATAA
TTCTTCGTTACTCATGCCATACATAAATGGATAGACGTCATATGAAGGTGTGCTTACAAGTGCTAATAATTCACCTGTTTGAGGGTGGATAGCAGTACCTGAGCCATAA
TTCTTCGTTACTCATGCCATACATAAATGGATAGACGTCATATGAAGGTGTGCTTACAAGTGCTAATAATTCACCTGTTTGAGGGTGGATAGCAGTACCTGAGCCATAA
TTCTTCGTTACTCATGCCATACATAAATGGATAGACGTCATATGAAGGTGTGCTTACAAGTGCTAATAATTCACCTGTTTGAGGGTGGATAGCAGTACCTGAGCCATAA
-----------------------------------------------------------------------------TGTTTGAGGGTGGATAGCAGTACCTGAGCCATAA
TTCTTCGTTACTCATGCCATACATAAATGGATAGACGTCATATGAAGGTGTGCTTACAAGTGCTAATAATTCACCTGTTTGAGGGTGGATAGCAGTACCTGAGCCATAA
S. haemolyticus
TTCTTCGTTACTCATGCCATACATAAATGGATAGACGTCATATGAAGGTGTGCTTACAAGTGCTAATAATTCACCTGTTTGAGGGTGGATAGCAGTACCTGAGCCATAA
TTCTTCGTTACTCATGCCATACATAAATGGATAGACGTCATATGAAGGTGTGCTTACAAGTGCTAATAATTCACCTGTTTGAGGGTGGATAGCAGTACCTGAGCCATAA
TTCTTCGTTACTCATGCCATACATAAATGGATAGACGTCATATGAAGGTGTGCTTACAAGTGCTAATAATTCACCTGTTTGAGGGTGGATAGCAGTACCTGAGCCATAA
TTCTTCGTTACTCATGCCATACATAAATGGATAGACGTCATATGAAGGTGTGCTTACAAGTGCTAATAATTCACCTGTTTGAGGGTGGATAGCAGTACCTGAGCCATAA
TTCTTCGTTACTCATGCCATACATAAATGGATAGACGTCATATGAAGGTGTGCTTACAAGTGCTAATAATTCACCTGTTTGAGGGTGGATAGCAGTACCTGAGCCATAA
S. hominis
TTCTTCGTTACTCATGCCATACATAAATGGATAGACGTCATATGAAGGTGTGCTTACAAGTGCTAATAATTCACCTGTTTGAGGGTGGATAGCAGTACCTGAGCCATAA
TTCTTCGTTACTCATGCCATACATAAATGGATAGACGTCATATGAAGGTGTGCTTACAAGTGCTAATAATTCACCTGTTTGAGGGTGGATAGCAGTACCTGAGCCATAA
TTCTTCGTTACTCATGCCATACATAAATGGATAGACGTCATATGAAGGTGTGCTTACAAGTGCTAATAATTCACCTGTTTGAGGGTGGATAGCAGTACCTGAGCCATAA
TTCTTCGTTACTCATGCCATACATAAATGGATAGACGTCATATGAAGGTGTGCTTACAAGTGCTAATAATTCACCTGTTTGAGGGTGGATAGCAGTACCTGAGCCATAA
TTCTTCGTTACTCATGCCATACATAAATGGATAGACGTCATATGAAGGTGTGCTTACAAGTGCTAATAATTCACCTGTTTGAGGGTGGATAGCAGTACCTGAGCCATAA
S. lugdunensis
TTCTTCGTTACTCATGCCATACATAAATGGATAGACGTCATATGAAGGTGTGCTTACAAGTGCTAATAATTCACCTGTTTGAGGGTGGATAGCAGTACCTGAGCCATAA
TTCTTCGTTACTCATGCCATACATAAATGGATAGACGTCATATGAAGGTGTGCTTACAAGTGCTAATAATTCACCTGTTTGAGGGTGGATAGCAGTACCTGAGCCATAA
TTCTTCGTTACTCATGCCATACATAAATGGATAGACGTCATATGAAGGTGTGCTTACAAGTGCTAATAATTCACCTGTTTGAGGGTGGATAGCAGTACCTGAGCCATAA
TTCTTCGTTACTCATGCCATACATAAATGGATAGACGTCATATGAAGGTGTGCTTACAAGTGCTAATAATTCACCTGTTTGAGGGTGGATAGCAGTACCTGAGCCATAA
TTCTTCGTTACTCATGCCATACATAAATGGATAGACGTCATATGAAGGTGTGCTTACAAGTGCTAATAATTCACCTGTTTGAGGGTGGATAGCAGTACCTGAGCCATAA

**b**

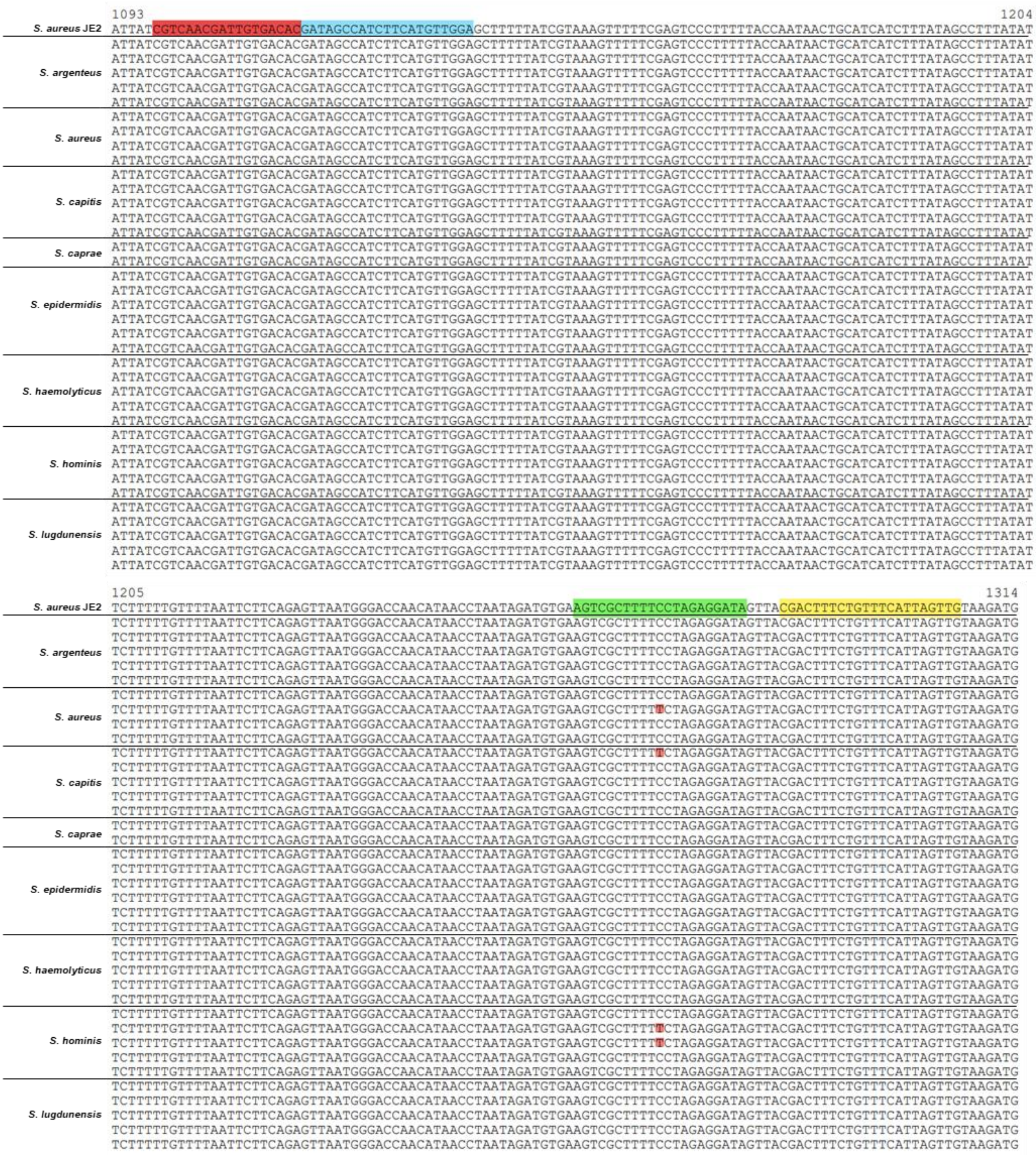


Figure S3. Alignment of ID4 (a) and ID16 (b) primer sets with *S. aureus* JE2 and comparison to other *mecA* sequences. F2 in blue; B2c in green and B3c in yellow. Mismatches are highlighted in red. Alignment from Benchling.

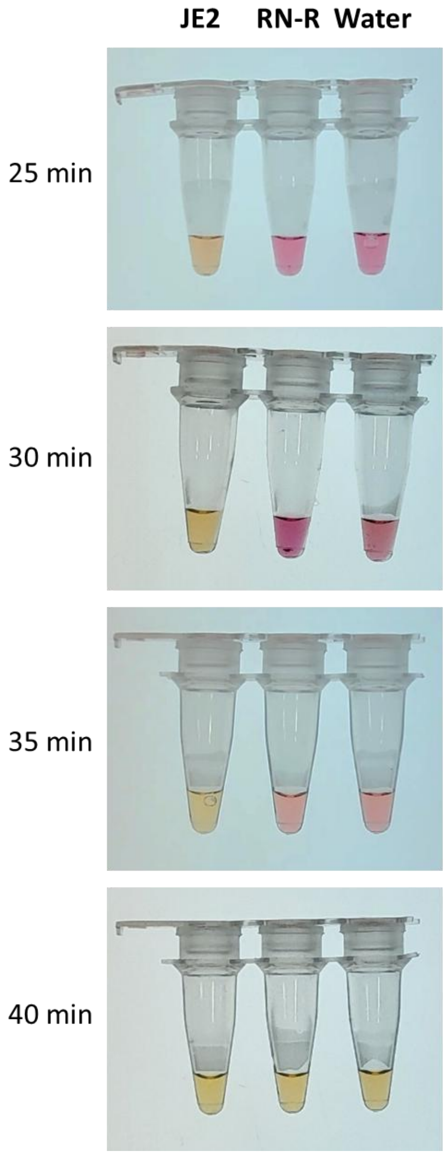


Figure S4. Colorimetric LAMP reaction performed with the newly designed ID16 primers. The pictures were taken at indicated reaction times.

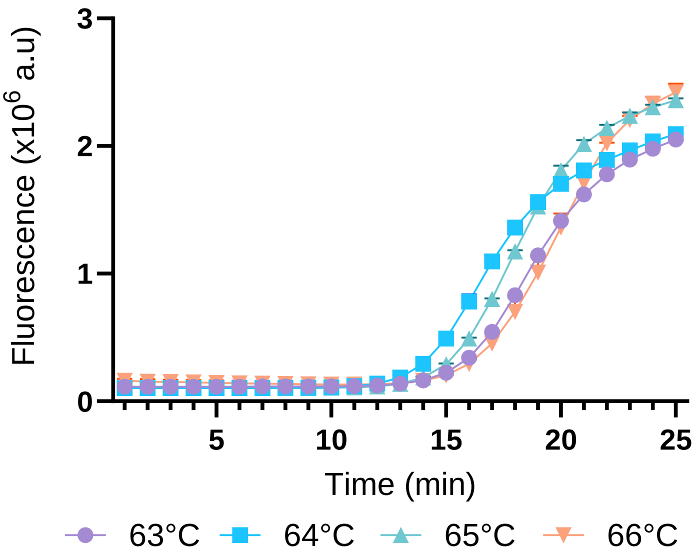


Figure S5. Fluorescent LAMP reaction at different temperatures. Performed using ID16 primer sets and 10 ng/µL of gDNA from JE2 strain.

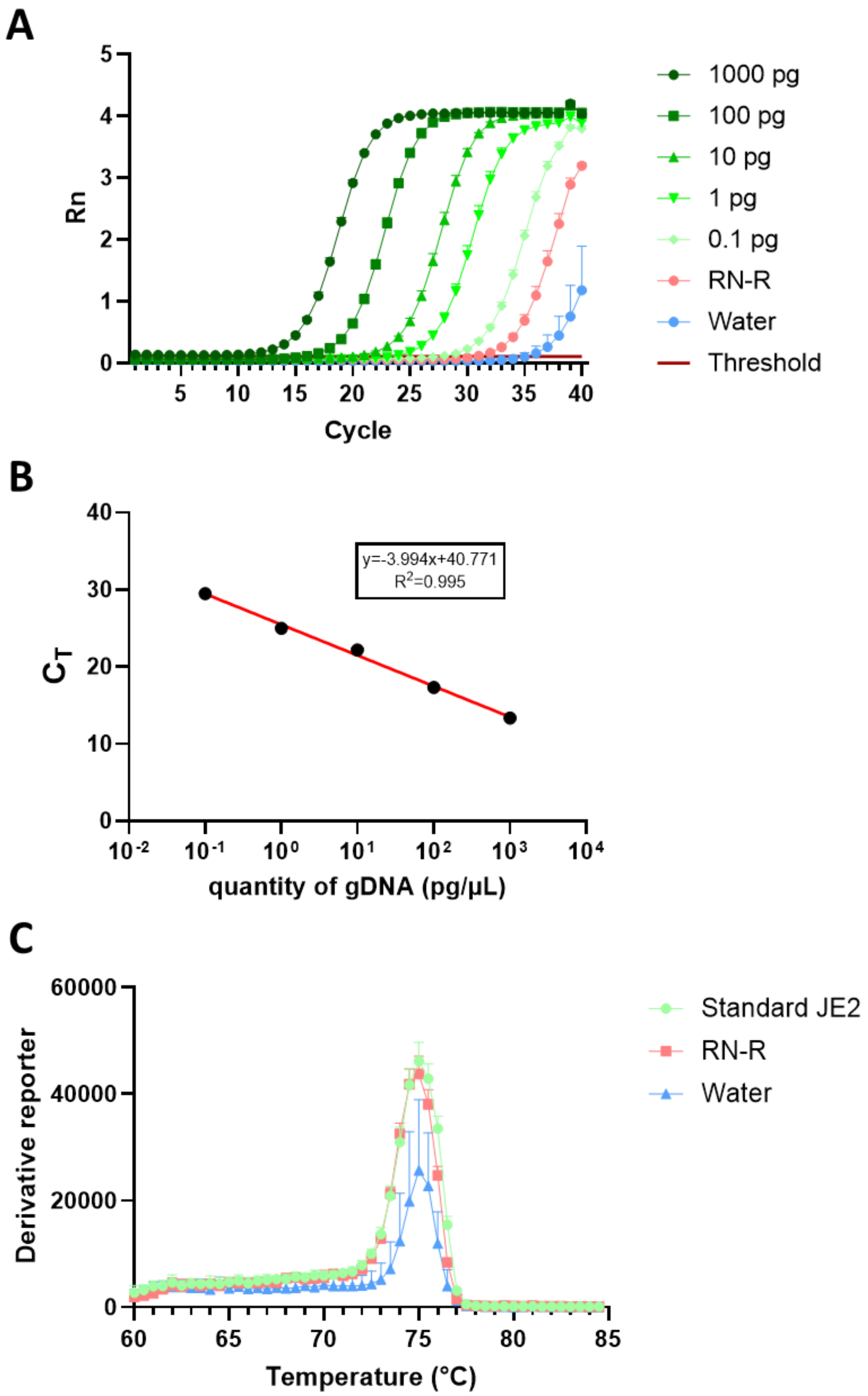


Figure S6. Sensitivity of the qPCR assay. A. Amplification plot: the amplification was performed using 10-fold serial dilution of gDNA from the JE2 strain carrying the *mecA* gene while 10 ng/µL of gDNA from RN4220 was used as the negative control. The mass of gDNA is expressed per µL of reaction. The threshold is automatically set by the qPCR software. B. Standard curve. C. Melting curve: the standard JE2 represent all dilutions.

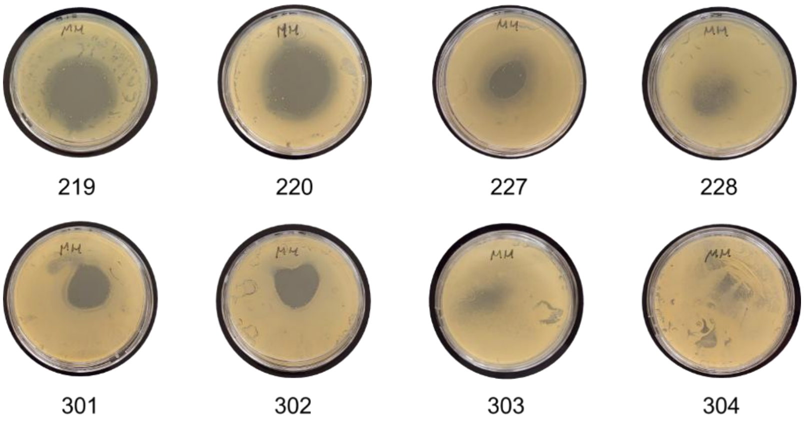


Figure S7. Methicillin susceptibility-test on Mueller-Hinton Agar plate. After spreading corresponding *S. aureus* cultures (foodborne strains named 219, 220, 227, and 228 for milk isolates, and 301-304 for meat isolates), 5 µg of methicillin were spotted in the center of plates. Pictures were taken after 24 h of incubation at 37°C.

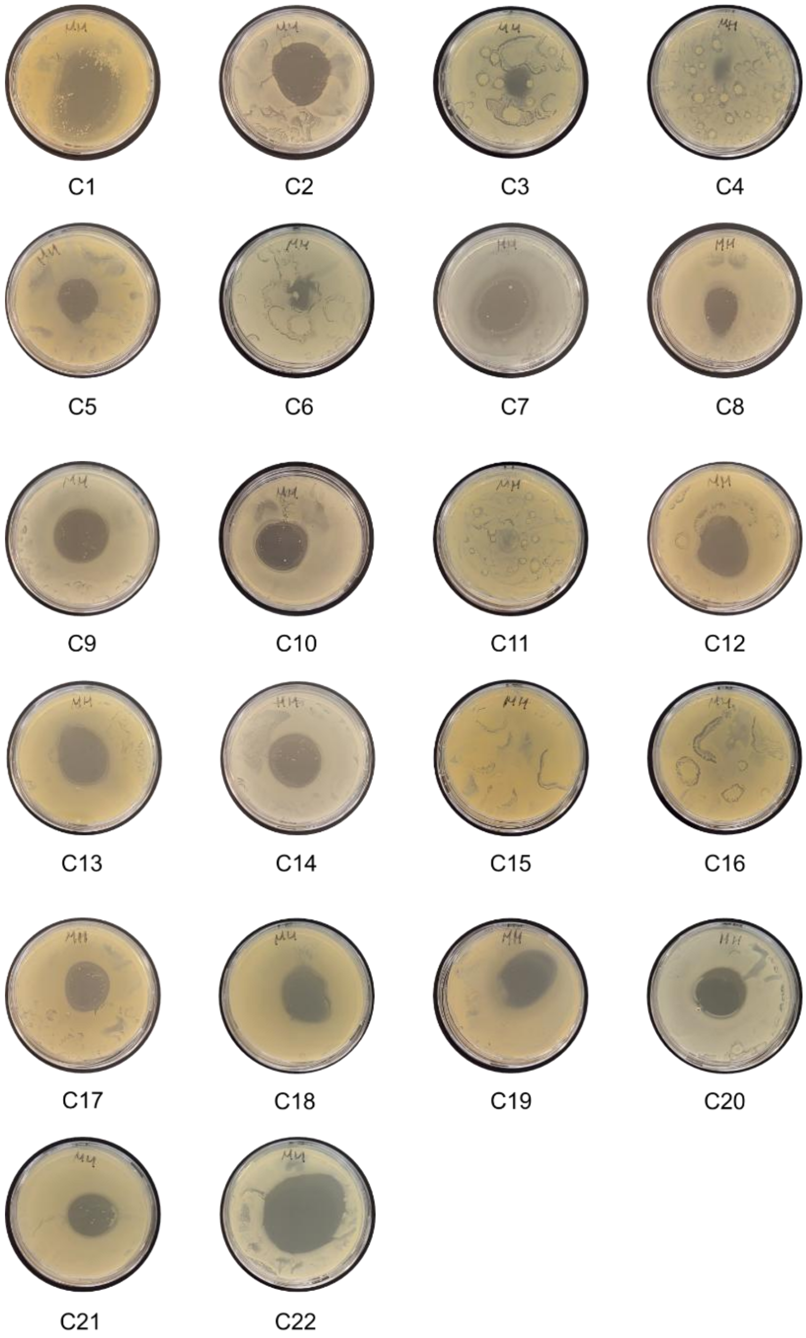


Figure S8. Methicillin susceptibility-test on Mueller-Hinton Agar plate. After spreading corresponding *S. aureus* cultures (clinical strains named C1-C22), 5 µg of methicillin were spotted in the center of plates. Pictures were taken after 24 h of incubation at 37°C.

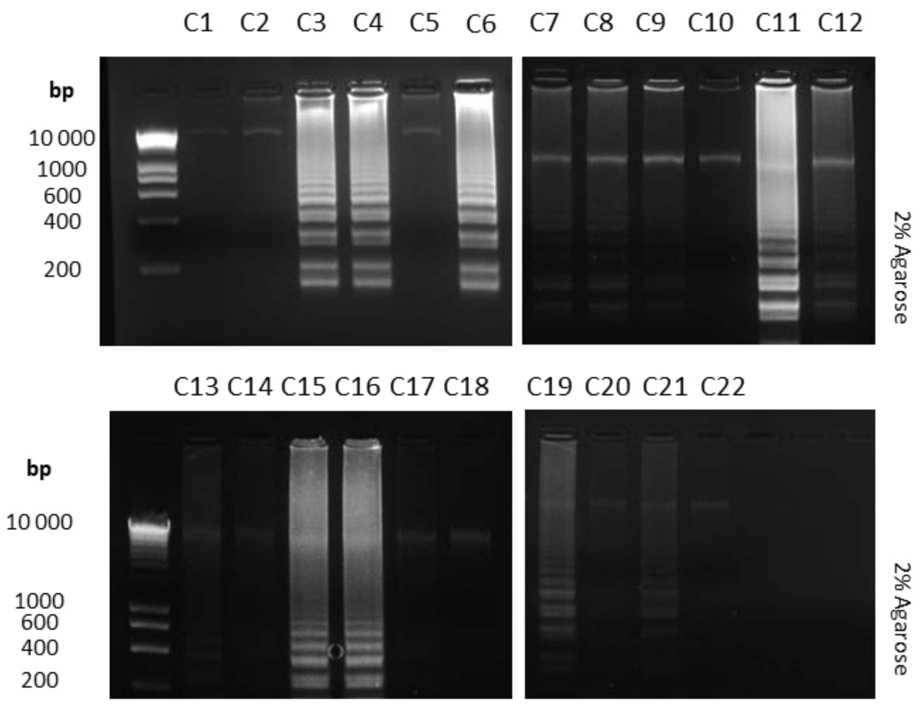


Figure S9. Validation of the colorimetric LAMP assay by agarose gel electrophoresis for detection *of S. aureus* clinical strains C1-C22, previously characterized for methicillin susceptibility as shown in Figure S8.
